# Relationships of gut microbiota, short-chain fatty acids, inflammation, and the gut barrier in Parkinson’s disease

**DOI:** 10.1101/2020.06.15.20131011

**Authors:** Velma T. E. Aho, Madelyn C. Houser, Pedro A. B. Pereira, Jianjun Chang, Knut Rudi, Lars Paulin, Vicki Hertzberg, Petri Auvinen, Malú G. Tansey, Filip Scheperjans

## Abstract

**Background:** Previous studies have reported that gut microbiota, permeability, short-chain fatty acids (SCFAs), and inflammation are altered in Parkinson’s disease (PD), but how these factors are linked and contribute to disease processes and symptoms remains uncertain.

**Objectives:** This study sought to compare and identify associations among these factors in PD patients and controls to elucidate their interrelations and links to clinical manifestations of PD.

**Methods:** Stool and plasma samples and clinical data were collected from 55 PD patients and 56 controls. Levels of stool SCFAs and stool and plasma inflammatory and permeability markers were compared between patients and controls and related to one another and to the gut microbiota.

**Results:** Calprotectin was increased and SCFAs decreased in stool in PD in a sex-dependent manner. Inflammatory markers in plasma and stool were neither intercorrelated nor strongly associated with SCFA levels. Age at PD onset was positively correlated with SCFAs and negatively correlated with CXCL8 and IL-1β in stool. Fecal zonulin correlated positively with fecal NGAL and negatively with PD motor and non-motor symptoms. Microbiota diversity and composition were linked to levels of stool SCFAs, inflammation, and zonulin. These relationships differed somewhat between patients and controls and by sex.

**Conclusions:** Intestinal inflammatory responses and reductions in fecal SCFAs occur in PD, are related to the microbiota and to disease onset, and are not reflected in plasma inflammatory profiles. Some of these relationships are PD- and sex-dependent. Alterations in microbiota-host interactions and links between intestinal inflammation and reduced SCFA levels and earlier PD onset warrant further investigation.

## Introduction

More than thirteen studies from three continents have established that the gut microbial communities of patients with Parkinson’s disease (PD) differ from those of healthy subjects.^1-3^ Most of these studies have focused on profiling the bacteria present in stool samples but have not linked these findings to functional consequences on microbiome-host interactions. Microbially-produced short-chain fatty acids (SCFAs) and regulation of immune responses and gut permeability are some of the proposed mechanisms by which gut microbes could impact brain health and function.^4, 5^

SCFAs, particularly acetic, propionic, and butyric acid, are the main end product of bacterial fiber fermentation in the gut.^6^ They have many effects on host physiology including acting as an energy source for colonocytes,^7, 8^ regulating the gut barrier,^9^ and influencing inflammatory responses.^10^ SCFAs have been suggested as key mediators in microbiota-gut-brain interactions.^11^ In a study contrasting fecal SCFA levels in PD patients and control subjects, decreased concentrations of SCFAs, particularly butyric acid, in PD patients were linked to microbiota alterations,^12^ and several bacterial taxa reportedly reduced in PD are SCFA producers.^2, 3, 13^

Inflammation is a widely recognized hallmark of PD. Increased levels of a variety of inflammatory molecules in brain and CSF^14-16^ as well as blood^17-19^ from PD patients have been documented. While the same specific cytokines, chemokines, and other inflammatory markers are not consistently implicated, IL-6, TNF, IL-1β, CRP, IL-10, CCL5, and IL-2 are among the most commonly dysregulated in the peripheral blood of PD patients.^18^ It has been suggested that this inflammation may result from intestinal barrier deficiency which could increase systemic exposure to inflammatory microbial products such as lipopolysaccharide (LPS), a component of bacterial cell walls.^5, 20^ Experiments measuring excretion of ingested sugars suggest increased permeability of the colon (but not the small intestine) in PD.^21-23^ Higher levels of zonulin and alpha-1-antitrypsin, indicators of gut permeability, have been found in PD patients’ stool relative to controls,^24^ although this difference has not been consistently observed.^25^ Additionally, increased detection of LPS^26^ and decreased LPS-binding protein (LBP)^21, 27^ in blood and plasma from PD patients suggest greater exposure of peripheral tissues and immune cells to LPS, supporting the existence of PD-related gut barrier dysfunction.

In recent years, there has been increasing recognition of a low-grade inflammatory state in the gut in PD. Studies have reported increased expression of genes encoding proinflammatory cytokines and chemokines in gut tissue from PD patients compared to controls,^22, 28^ and higher levels of IL-1α, IL-1β, CXCL8 (also known as IL-8), and CRP have been found in stool.^29^ The neutrophil-associated protein calprotectin is an indicator of gut inflammation in inflammatory bowel disease,^30^ and increased levels of calprotectin have been found in the stool of PD patients relative to controls.^24, 25^ Finally, higher numbers of CD3+ T cells and cells expressing the LPS receptor TLR4 have been identified in colon tissue from PD patients. When TLR4 was knocked out in a mouse model of PD pathology, symptoms were mitigated.^22^ Taken together, these findings support the hypothesis that intestinal bacteria and their metabolites along with inflammation and barrier dysfunction impact PD pathophysiology.

While there have been several studies evaluating inflammatory and gut permeability markers in PD, this study addresses the gap in knowledge regarding the interplay of these molecules in the blood and the gut and with the gut microbiota and SCFAs in PD patients and healthy control subjects.

## Materials and Methods

### Study subjects and sampling

The study was approved by the ethics committee of the Hospital District of Helsinki and Uusimaa. All participants gave informed consent. The study subjects were originally recruited for a pilot study of gut microbiota and PD.^31^ Samples for the present study were collected at a follow-up timepoint together with extensive clinical data, including questionnaires on diet, non-motor symptoms, and PD severity; full details have been published previously.^32^ Subjects collected stool samples at home into tubes with DNA stabilizer for microbiota analyses and tubes without preservatives for SCFA and immunological analyses. They kept the samples refrigerated until transport to the clinic (up to 3 days) for storage at −80°C. Venous blood samples were collected into EDTA tubes, centrifuged (10 min, 3000 rpm), and plasma was divided into aliquots and stored at -80°C. Samples were distributed to the laboratories by overnight shipping on dry ice. After excluding subjects with missing values for microbiota, acetic, butyric, or propionic acid, or most immune markers, the total number of subjects remaining for analysis was 111 (55 patients, 56 control subjects).

### Microbiota data

The microbiota data analyzed in this study have been published previously with a detailed description of the workflow^32^ and are available at the European Nucleotide Archive (accession number PRJEB27564). Briefly, stool samples were collected into PSP Spin Stool DNA Plus Kit tubes (STRATEC Molecular). DNA was extracted with the corresponding kit from the same manufacturer. The V3-V4 regions of the 16S rRNA gene were PCR-amplified and sequenced with Illumina MiSeq. Sequence quality control, Operational Taxonomic Unit (OTU) clustering, and taxonomical classification were performed with mothur.^33, 34^ Enterotypes were determined with an online tool.^35, 36^

### Short-chain fatty acid measurements

Detection and quantification of SCFAs in the stool samples were performed at the Norwegian University of Life Sciences using a Trace 1310 gas chromatograph (Thermo Fisher Scientific). Full details are available in Supplementary materials.

### Inflammatory and permeability marker measurements

Factors of interest in stool were measured using the Zonulin Stool ELISA (ALPCO 30-ZONHU- E01), LEGEND MAX(tm) Human MRP8/14 (Calprotectin) ELISA (Biolegend 439707), LEGEND MAX(tm) Human NGAL (Lipocalin-2) ELISA (Biolegend 443407), and the V-PLEX Proinflammatory Panel 1 Human (Meso Scale Discovery [MSD], Rockville, MD, K15049D) kits. The V-PLEX kit was also used to measure inflammatory mediators in plasma, as was the Human LBP Kit (MSD K151IYC). Stool and plasma samples were prepared, assays run, and results analyzed according to manufacturers’ protocols by the Emory Multiplexed Immunoassay Core. Full details are available in Supplementary materials.

### Statistical analyses

All statistical analyses were performed in R^37^ with packages including phyloseq,^38^ vegan,^39^ and DESeq2^40^ for microbial data comparisons. The complete R code is included as a supplementary file (Supplementary R Markdown), and full analysis details are available in Supplementary materials. For statistical comparisons between SCFAs/markers, clinical variables, enterotypes, and alpha diversity, we used the Kruskal-Wallis test, Wilcoxon rank sum test, or Pearson correlations depending on the types of variables. When multiple comparison corrections were included, we used the Benjamini & Hochberg false discovery rate (FDR), correcting SCFAs, stool markers, and plasma markers separately. Intercorrelated inflammatory markers were merged based on Principal Component Analysis.

## Results

### PD patients have higher calprotectin and lower SCFA levels in stool and lower CXCL8 levels in plasma compared to controls

Patient and control groups were similar with regard to basic demographics such as age, sex, and BMI; however, as we knew from previous analyses of the same subjects,^31, 32^ the groups differed regarding medications, medical history, and various symptom scores. A higher percentage of controls reported a history of stroke and use of medications for high blood pressure and cholesterol, and PD patients scored higher on scales of non-motor symptoms, gastrointestinal problems, and constipation (Table 1). Contrasting inflammatory markers and SCFAs between PD patients and control subjects, patients had lower levels of butyric and propionic acid and higher levels of calprotectin in their stool (Table S1) and lower levels of CXCL8 in plasma (Table S1). When the data were stratified by sex, the differences were particularly prominent for butyric acid in males and plasma CXCL8 and stool calprotectin in females (Fig. 1, Table S1).

**Table 1.** Demographic and Clinical Details of Subjects.

|  | **Control subjects** | **PD patients** | **p-value** |
| --- | --- | --- | --- |
| **Number of subjects** | 56 | 55 |  |
| **Sex (% Male)** | 48.21 | 52.73 | 0.706 |
| **Age at stool collection (mean ± SD)** | 66.38 ± 6.73 | 67.63 ± 5.21 | 0.293 |
| **Body Mass Index (mean ± SD)** | 26.7 ± 3.56 | 27.63 ± 4.74 | 0.261 |
| **History of TIA/ischemic stroke (%)** | 37.50 | 5.56 | < 0.001 |
| **Medication: ACE-inhibitor or AT1 antagonist (% yes)** | 48.21 | 29.09 | 0.051 |
| **Medication: calcium channel blocker (% yes)** | 19.64 | 5.45 | 0.042 |
| **Medication: statin (% yes)** | 48.21 | 18.18 | 0.001 |
| **Levodopa equivalent daily dose (LEDD mg; mean ± SD)** | 0 ± 0 | 608.61 ± 301.46 | < 0.001 |
| **Non-Motor Symptoms Scale (NMSS) total (mean ± SD)** | 7.09 ± 6.35 | 48.36 ± 36.55 | < 0.001 |
| **Rome III constipation subscore (items 9-15; mean ± SD)** | 2.46 ± 3.16 | 7.45 ± 5.22 | < 0.001 |
| **Rome III irritable bowel syndrome criteria fulfilled (%)** | 7.14 | 36.36 | < 0.001 |

SD - standard deviation, TIA - transient ischemic attack, ACE - Angiotensin-Converting Enzyme, AT1 - angiotensin II type 1

**Figure 1.**
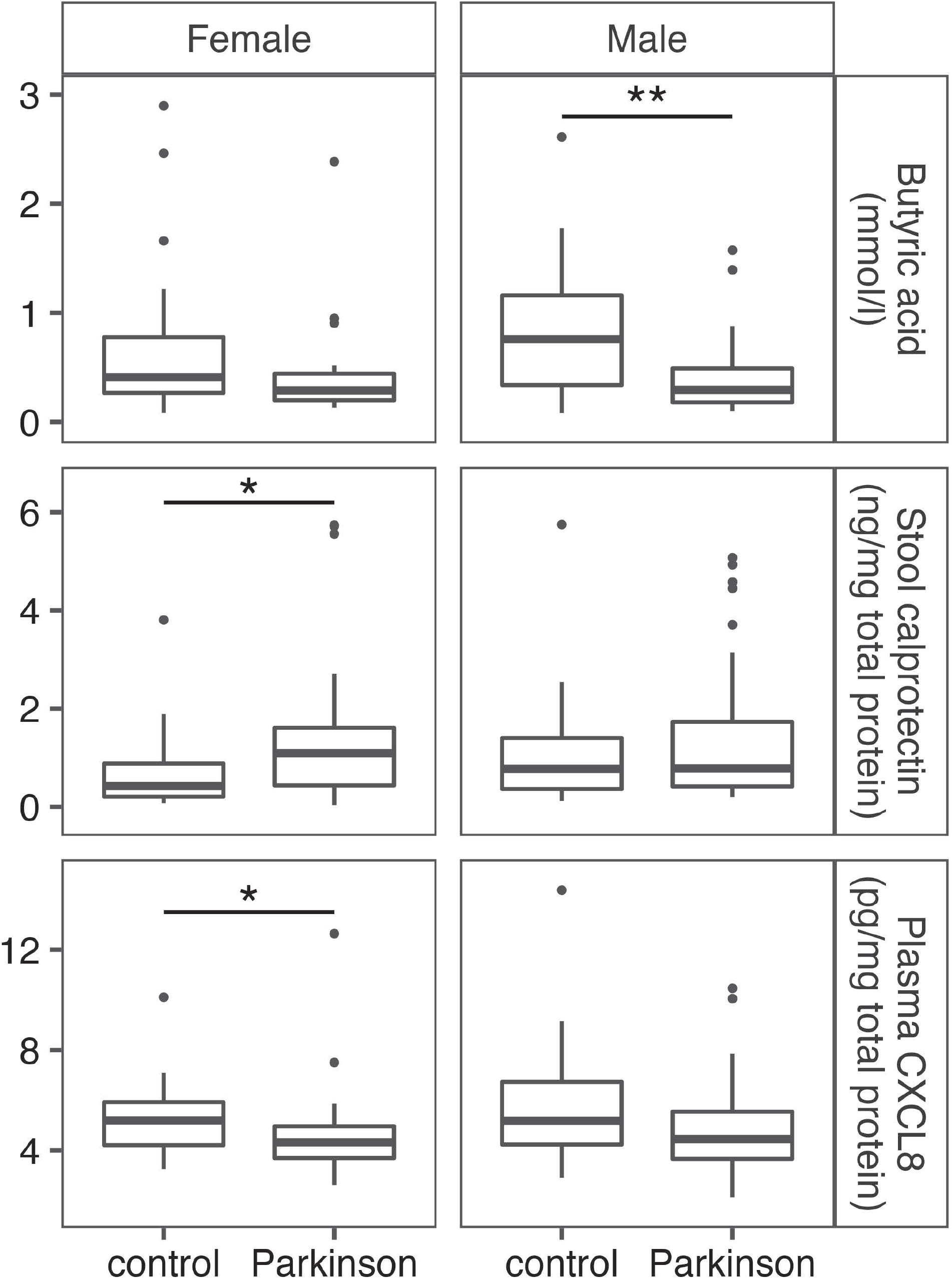
Differences in levels of stool butyric acid and calprotectin and plasma CXCL8 in male and female PD patients and control subjects.

### Inflammatory markers in plasma and stool are not correlated, but stool SCFAs, zonulin, and stool and plasma cytokines are related to PD onset and symptom severity

The inflammatory markers measured from the same sample material (plasma or stool) were largely intercorrelated, but we found no correlations between measurements of the same markers in stool and plasma (Fig. S1). PCAs for combining the most correlated markers resulted in one principal component (PC) for stool (summarizing IL-10, IL-12p70, IL-13, TNF, IL- 6, and IFNγ), and two plasma PCs (PC1: IL-10, IL-4, TNF; PC2: IL-1β, IL-2, IL-12p70, IL-13) (Supplementary R Markdown). In stool, the inflammatory markers neutrophil gelatinase-associated lipocalin (NGAL) and calprotectin and the gut permeability marker zonulin were significantly correlated (Fig. S1). In general, calprotectin and zonulin showed fewer correlations with other stool markers in the PD group as compared to the control group. Except for correlations with stool CXCL8 and IL-1β in the control group, none of the three most abundant SCFAs was significantly correlated with any of the markers in stool or plasma (Fig. S1).

Contrasting clinical variables against SCFAs and inflammatory markers suggested inverse correlations between stool SCFAs and several PD-related clinical variables such as the Non- Motor Symptoms Scale (NMSS) total score, the Rome III constipation/defecation subscore, and the Geriatric Depression Scale-15 (GDS15) (Figs. 2A, S2A). A similar pattern was seen for the Rome III constipation/defecation subscore, Rome III IBS criteria, and NMSS score and plasma CXCL8 (Figs. 2B, S2A). These correlations were significant for the entire cohort but not when evaluated only within PD patients. Butyric acid levels were correlated, however, with the age of onset for motor and non-motor symptoms of PD, while CXCL8 and IL-1β levels in stool were inversely correlated with the age of motor symptom onset (Fig. 2A, 2C, S2B). In PD patients, calprotectin levels were inversely linked to IBS symptoms, and stool zonulin was inversely correlated with variables related to PD severity, including the Unified Parkinson’s Disease Rating Scale (UPDRS) score, GDS15, and NMSS score (Fig. 2C, S2B). Symptom severity-related correlations remained significant after correcting for disease duration (Supplementary R Markdown).

**Figure 2.**
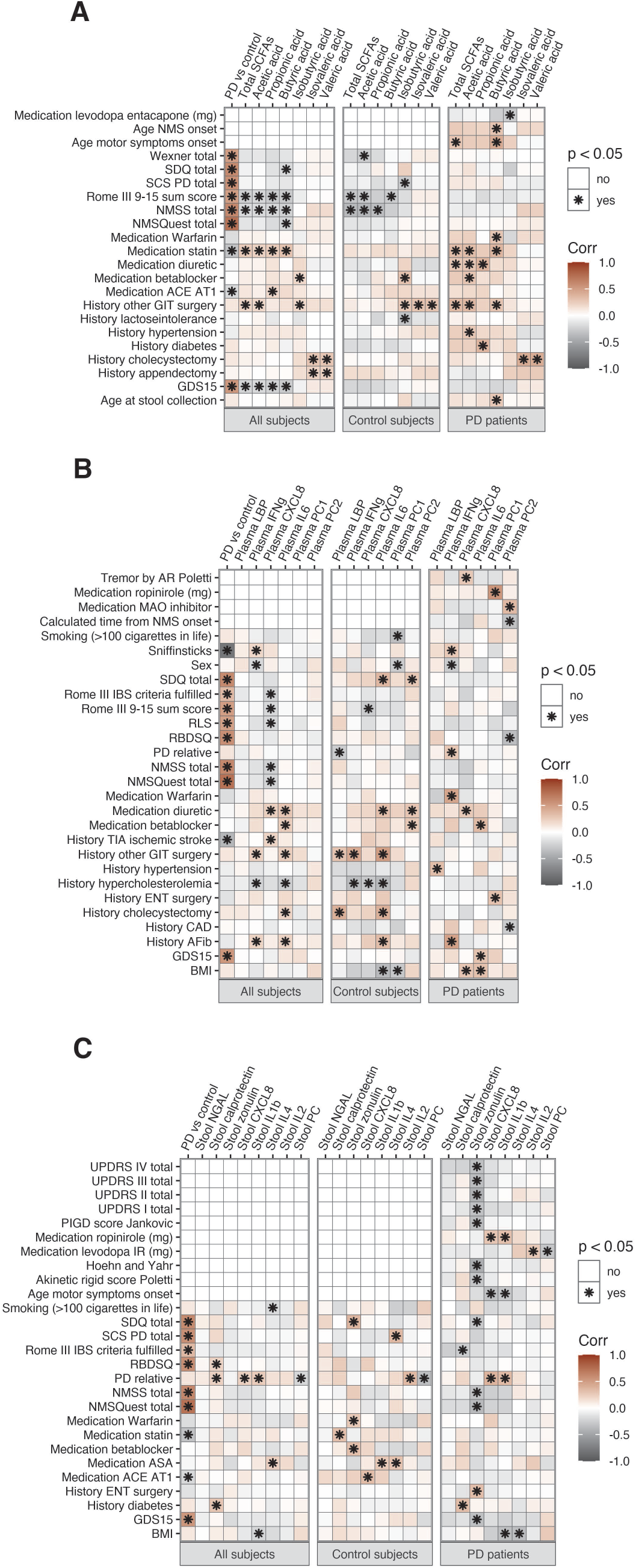
Correlations of SCFAs, inflammatory markers and clinical variables in full data and in only control subjects or PD patients.

### Microbial alpha diversity is inversely associated with stool inflammatory and permeability markers and SCFAs

Microbial alpha diversity indices, which indicate richness and evenness of bacterial taxa, were inversely correlated with total SCFAs, butyric and propionic acid, as well as with NGAL, calprotectin, zonulin, and IL-1β in stool (Fig. S3). Correlations between alpha diversity and SCFAs, NGAL, and zonulin were most evident in control subjects while correlations with calprotectin were most apparent in PD patients. In patients only, a significant inverse correlation between the Shannon diversity index and stool CXCL8 was also found (Fig. S3). Linear regression modeling corrected for sex and PD/control status suggested that the relationship between alpha diversity and stool inflammatory markers was independent of sex except for calprotectin, for which an inverse relationship was observed in females but not in males (Table S2A-C, Fig. 3A). There was a significant interaction between PD/control status and alpha diversity for propionic acid, showing an inverse correlation between microbial alpha diversity and propionic acid concentration in controls but not in PD patients (Fig. 3B). The total SCFAs variable showed a similar trend (Table S2B). The interaction in the propionic acid model remained significant after confounder correction (Table S2D).

**Figure 3.**
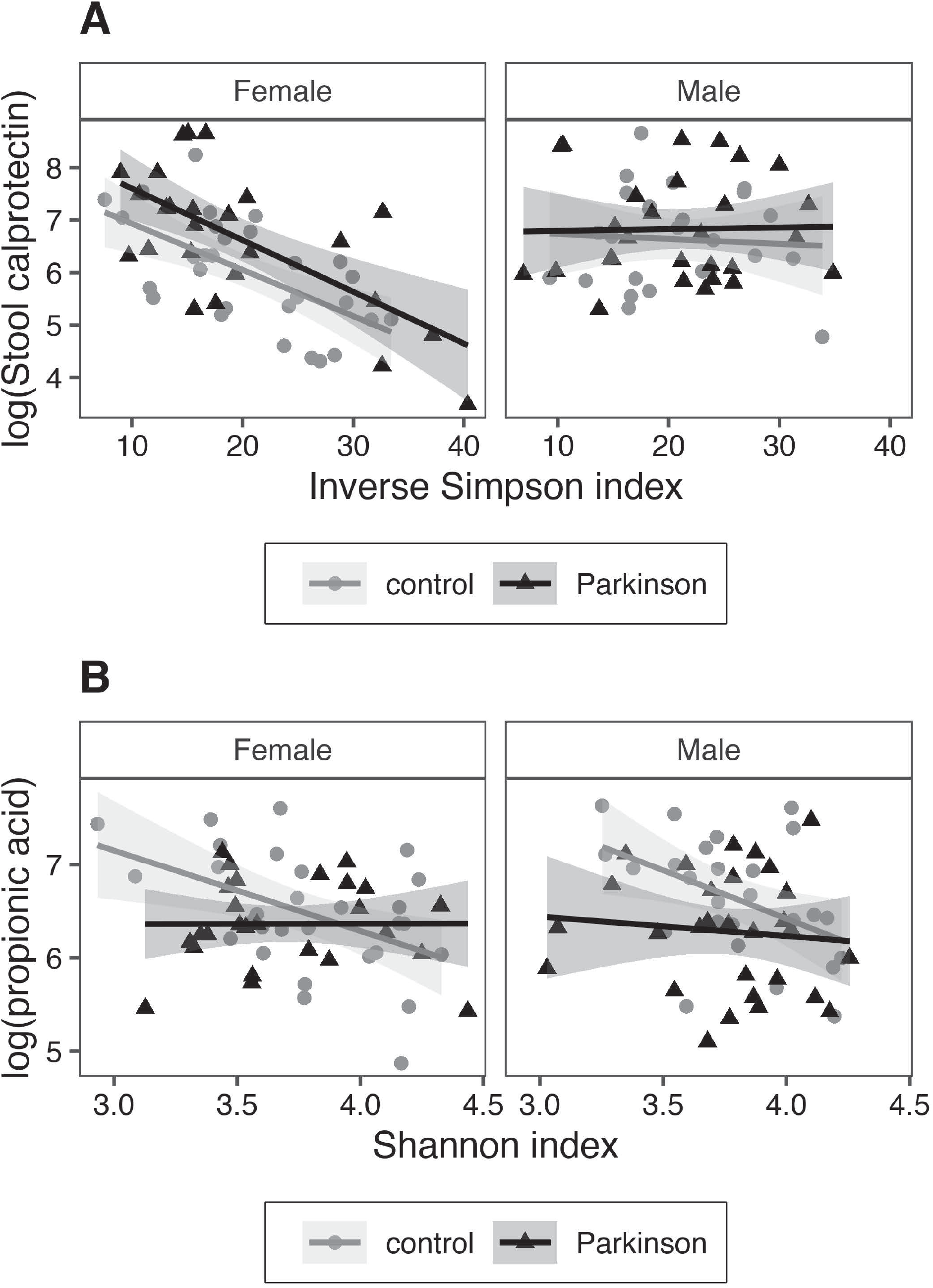
Linear modeling for alpha diversity showing effects of PD/control status and sex. **A)** Stool calprotectin and inverse Simpson index; **B)** Propionic acid and Shannon index.

### Microbial beta diversity is related to SCFA levels and inflammatory and permeability markers in stool

Contrasting community dissimilarity (beta diversity) and SCFAs or inflammatory markers, we found significant effects for total SCFAs, acetic, butyric, and propionic acid as well as stool NGAL, zonulin, IL2, and the stool marker PC in full data, with or without correction for PD/control status, sex, constipation, and BMI (Table S3A). The effect of calprotectin, while not statistically significant, was worth noting (*p* = 0.058 for full data, *p* = 0.079 when corrected for confounders). Control-only comparisons showed significant effects for SCFAs, NGAL, and zonulin; only butyric acid and zonulin were significant in PD-only comparisons (Table S3A, Fig. S4). These two variables also had a significant interaction with PD/control status when tested with the variable split into two categories by median (Table S3B).

### The *Prevotella* enterotype is associated with higher levels of butyric acid and lower levels of NGAL and zonulin in stool

One broad characterization method for microbiota composition is defining enterotypes based on certain indicator species in bacterial communities.^36^ In our subjects, stool zonulin, NGAL, propionic acid, and butyric acid had differences in concentrations between enterotypes (Table S4). Butyric acid levels were higher in PD patients with the *Prevotella* enterotype than the other two enterotypes, and control subjects with this enterotype had the lowest concentrations of NGAL and zonulin (Fig. S5). The findings regarding NGAL (full data and controls) and zonulin (controls only) remained significant after FDR correction (Table S4).

### Specific bacterial taxa are associated with levels of stool SCFAs and inflammatory and permeability markers

Guided by the associations with beta diversity, we explored the associations of specific bacterial taxa with the three most abundant SCFAs and with stool NGAL, calprotectin, and zonulin. The abundances of *Butyricicoccus, Clostridium sensu stricto*, and *Roseburia* were positively correlated with levels of SCFAs while the abundances of *Akkermansia, Escherichia/Shigella, Flavonifractor, Intestinimonas, Phascolarctobacterium*, and *Sporobacter* decreased with increasing SCFA concentrations (Fig. 4, S6, Table S5A). Similar patterns could be seen for the parent families of these taxa, such as a positive association between butyric acid and *Lachnospiraceae* and a negative one for *Enterobacteriaceae* and *Verrucomicrobiaceae* (Table S5A). A positive association between SCFAs and *Bacteroides* was observed only in controls, while a negative association with *Bifidobacterium* was observed only in PD patients (Fig. 4, S6, Table S5A).

**Figure 4.**
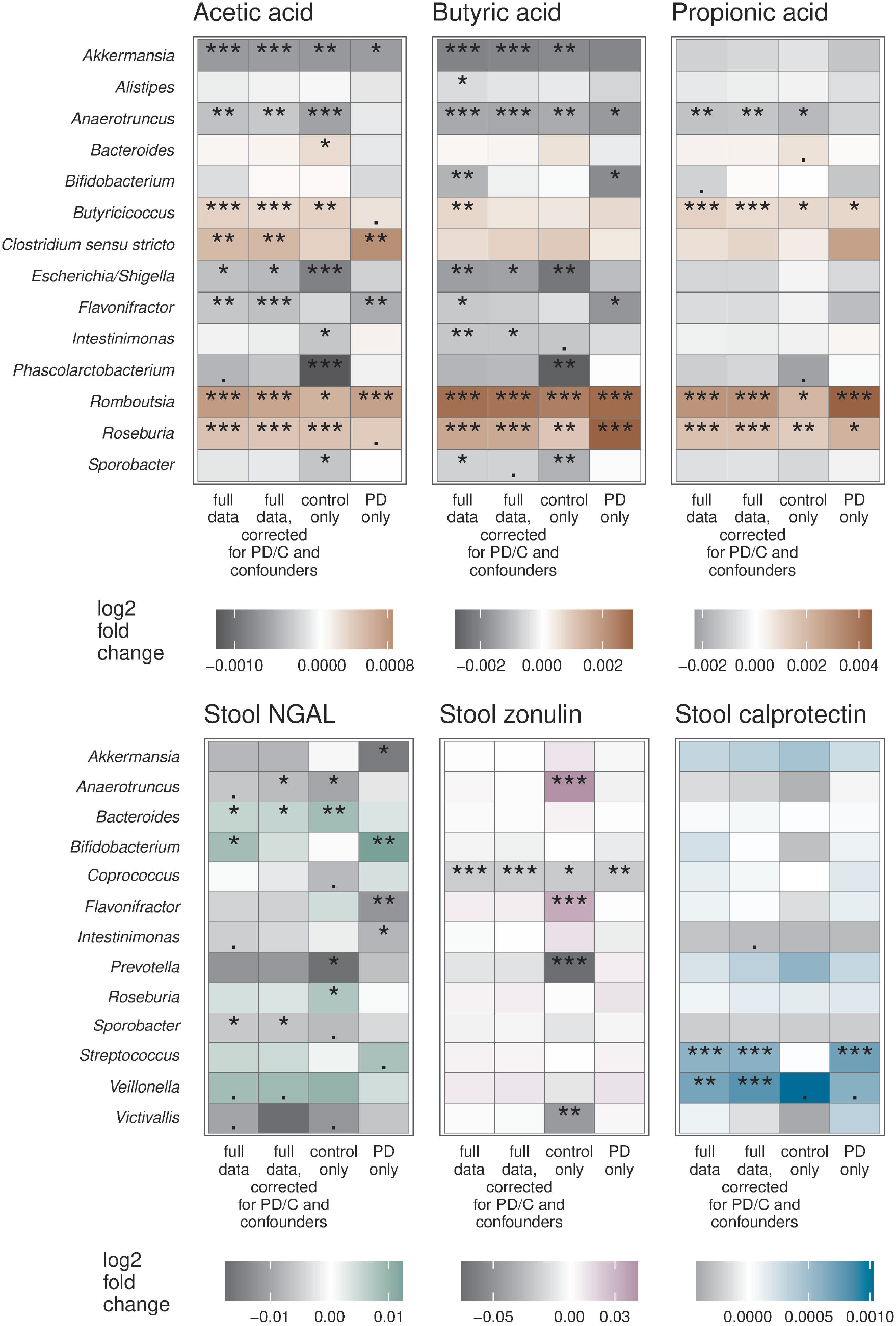
Associations of bacterial genera, SCFAs, and stool inflammatory markers. Confounders included in confounder-corrected models were Rome III 9-15 sum score and sex. Significance level is marked as follows: *** : p ≤ 0.001 / ** : p ≤ 0.01 / * : p ≤ 0.05 /. : 0.1 > p > 0.05.

Microbiota associations with the stool markers differed between PD patients and control subjects. Stool NGAL levels were positively associated with *Bifidobacterium* in PD but with *Bacteroides* and *Roseburia* in controls (Fig. 4, S7A, Table S5B). Relative abundance of *Prevotella* was negatively associated with both NGAL and zonulin in controls only (Fig. 4, S7A- B, Table S5B). The only microbiota-immune association found to be significant in both PD patients and controls was the negative relationship between stool zonulin and the genus *Coprococcus* (Fig. 4, S7B, Table S5B), a taxon known to have a strong influence on gut permeability^41^ and stool zonulin levels.^42^ Other associations with low-abundance taxa were statistically significant but were driven by a small number of data points, precluding confident interpretation (Fig. S7).

## Discussion

Numerous studies have reported abnormalities in the gut microbiome, levels of SCFAs, peripheral immune responses, and intestinal permeability in PD; however, this study provides for the first time an integrative analysis of these factors in the same subjects.

Efforts have been made to identify biomarkers of PD in both blood and stool, but the relationship between the levels of inflammatory markers in the gut and in circulation in this disorder had not been elucidated. We determined that levels of inflammation-associated molecules in the plasma and in the stool were highly correlated within each sample type. We found essentially no correlation, however, between levels of inflammatory markers in plasma and stool. This indicates that systemic and gut immune responses are distinct and, thus, measurements from one site are unlikely to represent the nature of the responses in the other.

In the plasma, we found a significant decrease in levels of the chemokine CXCL8 (also known as IL-8) in female PD patients as well as an inverse correlation between CXCL8 and non-motor symptom severity scores in the full cohort. CXCL8 is the primary chemoattractant for neutrophils and is commonly produced in inflammatory responses to stimulate and recruit these phagocytic cells. A previous study has reported a positive correlation between CXCL8 levels in the serum and the degree of PD-related disability,^43^ rendering our findings somewhat unexpected. A possible explanation is that plasma CXCL8 levels in this cohort are significantly higher in subjects with a history of stroke (Fig. 2), the majority of whom are controls.

In stool, we documented lower levels of SCFAs and higher levels of calprotectin in PD patients. This is consistent with other reports of SCFA and SCFA-producing bacteria reductions^3, 12, 13, 19, 22, 31^ as well as increased stool calprotectin^24, 25^ in PD. While SCFAs and calprotectin are beginning to emerge as some of the most reliable indicators of a dysregulated and inflammatory gut environment in PD, our results suggest that these associations may be sexually dimorphic. Levels of butyric acid specifically were correlated with the age of onset for both motor and non-motor PD symptoms even after accounting for disease duration, raising the possibility that higher butyric acid levels could be protective and delay disease onset. We also found inverse correlations between stool SCFA levels and non-motor, particularly gastrointestinal and depressive, symptoms. This correlation was not significant in just the PD subset of the cohort, however, which could be due to a lack of statistical power or to a difference between the control and patient groups.

Though SCFAs are known to have immunomodulatory and barrier-promoting properties, we found no significant correlations between the most abundant stool SCFAs and immune- or permeability-related factors in stool or plasma. While it is possible that SCFA levels may directly impact immunological pathways not assessed in this study, the lack of clear association between SCFA production and inflammation likely evinces the complexity of the relationship between these physiological processes. Both pro- and anti-inflammatory activities of SCFAs have been documented,^44, 45^ and dietary supplementation of SCFAs produces variable effects on inflammation which appear to be strongly influenced by the range of SCFA concentrations involved.^46^ Further investigation of the effects of SCFAs on immune responses in the gut, the blood, and the central nervous system in the context of PD pathology is needed.

NGAL, an epithelium-derived antimicrobial glycoprotein overexpressed in inflammatory conditions,^47^ has been found at higher levels in the plasma of PD patients compared to controls.^43^ To the best of our knowledge, this comparison had not been performed in stool, and we found no significant differences in fecal NGAL levels between patients and controls. We also found no significant indications of greater intestinal permeability in this cohort of PD patients as measured by stool zonulin or plasma LBP, though other studies have.^21, 24, 27^ On the contrary, we found that higher zonulin levels were associated with less severe clinical manifestations of PD. This could be a reflection of the diversity of motor and non-motor symptom subtypes in PD.^48^ Longitudinal studies assessing intestinal permeability beginning in the earliest stages of PD would be beneficial to determine its impact on disease symptoms and progression.

It has been proposed that intestinal inflammation in PD is related to gut permeability and alterations in microbiota composition and can contribute to disease pathogenesis.^5^ In this study, we found that levels of fecal NGAL were positively correlated with fecal zonulin, supporting a relationship between intestinal inflammation and permeability. Levels of inflammatory and permeability markers were also associated with the stool microbiota. Stool NGAL, calprotectin, zonulin, and CXCL8 decreased with increasing alpha diversity, consistent with the concept that an inflamed gut environment exerts selective pressure on the microbiota. NGAL, IL-2, the stool marker PC, and zonulin were also significantly associated with alterations in beta diversity, and the association with calprotectin nearly reached the threshold for significance. Furthermore, the inflammatory factors CXCL8 and IL-1β in stool, which were found to be increased in a different cohort of PD patients,^29^ were negatively correlated with the age of PD motor symptom onset in this study, supporting the involvement of gut inflammation in the development of PD pathology.

A notable finding in this study is that the relationships between gut microbes and inflammatory factors and between microbes and their metabolites such as SCFAs in stool differ between PD patients and controls. In control subjects only, SCFAs were inversely associated with alpha diversity, and SCFA associations with beta diversity were stronger in controls than PD patients. Conversely, levels of butyric acid in stool differed significantly by enterotype only among PD patients. Relationships between SCFAs and individual taxa were largely concordant in both subject groups, including positive correlations between stool SCFAs and well-known SCFA producers like *Butyricicoccus* and *Roseburia* and a negative correlation with *Akkermansia*, which SCFAs are known to inhibit.^49^ On the other hand, a positive correlation between SCFAs and relative abundance of *Bacteroides* was found only in controls while a negative correlation with *Bifidobacterium* was found only in PD patients.

These discrepancies could reflect differences in microbiota composition between patients and controls; *Bacteroides* is more abundant in controls, and *Bifidobacterium* is more abundant in PD patients in this cohort,^32^ which could increase their functional impacts within their respective microbial communities. This could explain the positive association between SCFAs and *Bacteroides*, a producer of SCFAs, in controls, but not necessarily the inverse relationship between *Bifidobacterium* and butyric acid in PD patients. Increases in *Bifidobacterium* are considered butyrogenic due to cross-feeding of butyrate-producing bacteria,^50^ but the opposite association was found in our PD cohort, suggesting an altered function or strain profile of the *Bifidobacterium* genus in the metabolic network of the PD microbiota. Further evidence that *Bifidobacterium* may not be performing a homeostatic function in PD is its positive correlation with the inflammatory marker NGAL in patients only.

PD patients in this cohort had reduced abundance of *Prevotella*,^32^ and significant links between lower levels of fecal zonulin and NGAL and *Prevotella* abundance and enterotype were observed only in the control group. Visual inspection of the data in Figs. S5 and S7 suggests that a similar trend in the relationship between NGAL and *Prevotella* exists in the PD group, and differences in the strength of the association in patients and controls may be influenced by the lower abundance of *Prevotella* in PD. Whether the relationship between *Prevotella* and zonulin in PD differs from that in controls is less clear, and, as this taxon is frequently reported to be impacted in PD,^19, 31, 32, 51-53^ future studies evaluating its influence on intestinal permeability in patients are warranted.

It has been established that alterations in the composition of the gut microbiota and intestinal immune responses occur in PD, but our findings suggest that the activities of particular bacteria and the nature of their interactions with the host immune system may be changing as well. Future longitudinal microbiome studies using multi-omics approaches with higher taxonomic resolution combined with detailed pheno- and genotyping of subjects will provide a thorough understanding of how gut microbes and their metabolites are interacting with the host and impacting the etiology, symptoms, and progression of PD. This has the potential to reveal new targets along the gut-brain axis for more effective disease-modifying treatment of this disorder.

## Data Availability

All statistical analyses were performed in R37 with packages including phyloseq,38 vegan,39 and DESeq240 for microbial data comparisons. The complete R code is included as a supplementary file (Supplementary R Markdown), and full analysis details are available in Supplementary materials.

## Acknowledgements

This study was supported in part by the Emory Multiplexed Immunoassay Core (EMIC), which is subsidized by the Emory University School of Medicine and is one of the Emory Integrated Core Facilities. The content of this paper is solely the responsibility of the authors and does not necessarily reflect the official views of the National Institutes of Health.

## Authors’ Roles

1. Research project: A. Conception, B. Organization, C. Execution
2. Statistical Analysis: A. Design, B. Execution, C. Review and Critique
3. Manuscript: A. Writing of the first draft, B. Review and Critique

Velma T. E. Aho: 1C, 2A, 2B, 2C, 3A, 3B

Madelyn C. Houser: 1A, 1B, 1C, 2A, 2C, 3A, 3B

Pedro A. B. Pereira: 1B, 1C, 2C, 3B

Jianjun Chang: 1B, 1C, 3B

Knut Rudi: 1C, 3B

Lars Paulin: 1B, 1C, 3B

Vicki Hertzberg: 2A, 2C, 3B

Petri Auvinen: 1A, 1B, 1C, 2C, 3B

Malú G. Tansey: 1A, 1B, 2A, 2C, 3B

Filip Scheperjans: 1A, 1B, 1C, 2C, 3B

## Financial Disclosures of all authors (for the preceding 12 months)

Rudi: none

Hertzberg: NIH (P50ES02607, P30NR018090, R01LM013323, HHSN272201400004C), EPA(83615301), ALS Association (20-IIA-524, also funds Madelyn Houser), CDC/NIOSH (R01OH011782)

Auvinen: Päivikki and Sakari Sohlberg foundation

Tansey: NIH (1R01NS092122, 1RF1AG051514, 1RF1AG057247, also funded Madelyn Houser and Jianjun Chang), Michael J. Fox Foundation for Parkinson’s Research, and the Parkinson’s Foundation, National Center for Georgia Clinical & Translational Science Alliance of the National Institutes of Health (UL1TR002378). Advisor to INmune Bio, Longevity Biotech, Prevail Therapeutics, and Weston Garfield Foundation. MGT has patents issued (US Pat. Nos.

7144987B1 and 7244823B2) and pending (US20150239951, WO2019067789, 62/901698, see efiling Ack37193677, 62/905747, see efilingAck37274773) for co-invention of DN-TNFs. Scheperjans: Grants from Academy of Finland, Hospital District of Helsinki and Uusimaa, The European Commission, Renishaw; consulting fees and honoraries from Abbvie, Orion, LivaNova, Axial Biotherapeutics, NeuroInnovation; non-financial support from NordicInfu Care, Herantis Pharma, Global Kinetics Corporation, Abbott. Founder and CEO of NeuroInnovation Oy and NeuroBiome Ltd.

VTEA, PABP, LP, PA, and FS have patents FI127671B & US10139408B2 issued and patents US16/186,663 & EP3149205 pending that are assigned to NeuroInnovation Oy and licensed to NeuroBiome Ltd.

## Supplement

Supplementary Methods, with extended descriptions of measurements for SCFAs and inflammatory and permeability markers, and details of statistical analyses.

Supplementary R Markdown, showing the full R code used for the analyses.

Figure S1. Correlations for variables of interest in full data and subsets. **A)** Full data; **B)** Control subjects; **C)** PD patients. * : p < 0.05.

Figure S2. Scatterplots for significantly correlated clinical variables and SCFAs or inflammatory or permeability markers. **A)** Variables measured in all subjects; **B)** Variables measured only in PD patients.

Figure S3. Correlations of inverse Simpson and Shannon diversity for SCFAs and inflammatory and permeability markers. Significance level is marked as follows: *** : p ≤ 0.001 / ** : p ≤ 0.01 / * : p ≤ 0.05 /. : 0.1 > p > 0.05.

Figure S4. NMDS ordination plots for beta diversity, PD/control status, and the stool markers and SCFAs associated with the most notable beta diversity difference. Variables were split into two categories by median.

Figure S5. Butyric acid, stool NGAL, and zonulin in different enterotypes among PD patients and control subjects

Figure S6. Scatter plots of differentially abundant genera for SCFAs

Figure S7. Scatter plots of the differentially abundant genera for stool inflammatory and permeability markers. **A)** NGAL; **B)** Zonulin; **C)** Calprotectin.

Table S1. Marker and SCFA concentrations by PD/control status and sex

Table S2. Linear regression for alpha diversity, inflammatory and permeability markers/SCFAs, and PD/control status. **A)** Model: log(variable) ∼ sex + PD/control + Diversity; **B)** Model: log(variable) ∼ sex + PD/control * Diversity; **C)** Stool calprotectin and additional confounders; **D)** Propionic acid and additional confounders.

Table S3. Beta diversity, inflammatory and permeability markers/SCFAs and PD status. **A)** Without interactions; **B)** With interaction (model: community dissimilarity ∼ PD/control * variable).

Table S4. Differences in inflammatory and permeability marker/SCFA levels between enterotypes

Table S5. Differentially abundant taxa for variables of interest. Table includes the results for families and genera that had a multiple comparison corrected p < 0.05 in at least one comparison for **A)** Acetic, propionic, and butyric acid or **B)** Stool NGAL, zonulin, and calprotectin.

